# Vaccine-induced antibody level predicts the clinical course of breakthrough infection of COVID-19 caused by delta and omicron variants: a prospective observational cohort study

**DOI:** 10.1101/2022.03.09.22272171

**Authors:** Min Hyung Kim, Yooju Nam, Nak-Hoon Son, Namwoo Heo, Bongyoung Kim, Eawha Kang, Areum Shin, Andrew Jihoon Yang, Yoon Soo Park, Heejung Kim, Taeyoung Kyong, Yong Chan Kim

## Abstract

**Background:** Omicron variant viruses spread rapidly, even in individuals with high vaccination rates. This study aimed to determine the utility of the antibody against the spike protein level as a predictor of the disease course of COVID-19 in vaccinated patients.

**Methods:** Between 11 December 2021 and 10 February 2022, we performed a prospective observational cohort study in South Korea, which included patients infected with delta –and –omicron variants. Multivariable logistic regression analysis to determine the association between antibody levels and the outcomes was conducted.The relationship between antibody levels and cycle threshold (Ct) values was confirmed using a generalised linear model.

**Results:** From 106 vaccinated patients (39 delta and 67 omicron), the geometric mean titres of antibodies in patients withfever (≥37.5 °C), hypoxia (≤94% of SpO_2_), pneumonia, C-reactive protein (CRP) elevation (>8 mg/L), or lymphopenia (<1,100 cells/μL) were 1,201.5 U/mL, 98.8 U/mL, 774.1 U/mL, 1,335.1 U/mL, and 1,032.2 U/mL, respectively. Increased antibody levels were associated with a decrease in the fever occurrence (adjusted odds ratio [aOR], 0.23; 95% confidence interval [CI], 0.12–0.51), hypoxia (aOR, 0.23; 95% CI, 0.08–0.7), CRP elevation (aOR, 0.52; 95% CI, 0.29–0.0.94), and lymphopenia (aOR, 0.57; 95% CI, 0.33–0.98). Ct values showed a positive correlation between antibody levels (P =0.02).

**Conclusion:** Antibody levels are predictive of the clinical course of COVID-19 in vaccinated patients with delta and omicron variant infections. Our data highlight the need for concentrated efforts to monitor patients with SARS-CoV-2 infection who are at risk of low antibody levels.

**Summary:** In this prospective observation cohort study, antibody level predicts clinical course of breakthrough infection of COVID-19. Fever (aOR 0.23[0.12-0.51], hypoxia (aOR 0.23[0.08-0.7]), CRP elevation(aOR 0.52[0.29-0.0.94] and lymphopenia (aOR 0.57[0.33-0.98]) were inversely correlated with antibody levels.

## Introduction

Since the omicron variant was first confirmed on 24 November 2021, it has spread rapidly worldwide[1]. As a result, global cases of severe acute respiratory syndrome coronavirus 2 (SARS-CoV-2) infection have increased at an unprecedented rate[2]. The omicron variant can spread easily in individuals even if they have completed their vaccination course. Several mutations in the omicron variant are expected to enable the virus to evade the immune system established by vaccination, resulting in increased infectivity[3,4]. In addition, a decrease in immunity elicited by vaccines over time, namely waning immunity, may play an important role in the omicron variant’s spread[5]. However, vaccines remain effective in protecting against severe diseases caused by the omicron variant, although the effectiveness may have decreased in the omicron variant rather than in previous variants[6]. Understanding the extent of vaccine effectiveness on clinical protection in patients with breakthrough infection caused by the omicron variant is needed to take measures to minimise the damage from the current pandemic.

In South Korea, owing to quarantine and isolation guidelines, it took more time for the omicron variant to become the dominant strain than in other countries. According to several reports, the omicron variant virus presumably has weakened virulence, which is related to reduced severity, hospitalisation, and mortality[7,8]. However, the recent explosive increase in the number of omicron variant infections has increased the total number of deaths, and the omicron’s spread has not slowed as of 28 February 2022. In this situation, to efficiently use limited medical resources, it is important to predict which patients will go through an unfavourable clinical course and put concentrated efforts into them.

We performed a prospective cohort study involving patients with delta and omicron variant infections who were admitted to an institution in South Korea. This study aimed to determine the vaccine’s effect on the clinical course of delta and omicron variant infections. Furthermore, we evaluated the usefulness of the antibody level to spike protein as a predictor of the disease course of COVID-19 in vaccinated patients.

## Methods

### Study design and participants

We performed a prospective observational cohort study involving SARS-CoV-2 confirmed adult patients (age, >19 years) who were admitted to the Yongin Severance Hospital. All SARS-CoV-2 cases were confirmed by polymerase chain reaction (PCR) tests and then notified to a local public health centre in South Korea. Patients with specified symptoms or conditions were asked to be hospitalised for monitoring and treatment, and they were assigned to appropriate hospitals according to their severity of COVID-19 if they were willing to do so. Symptoms and conditions for which hospitalisation was considered and classification of severity are described in appendix p2-3 (Supplementary table 1 and 2). Yongin Severance Hospital has been in charge of hospitalisation for SARS-CoV-2 cases with mild to moderate severity.

This study enrolled participants from 11 December 2021 to 10 February 2022, which were designated to include two different waves of COVID-19 by delta and omicron variants in South Korea. Based on the national data of COVID-19, almost all SARS-CoV-2 cases were caused by the delta variant until the start of study enrolment. However, the omicron variant overtook the delta variant in domestic SARS-CoV-2 cases from the third week of January 2022, and its detection rate exceeded 90% in the first week of February 2022 (Supplementary figure 1). Patients who agreed to undergo PCR tests for the SARS-CoV-2 variant type and anti-SARS-CoV-2 antibody tests were eligible for enrolment in this study. Only participants with confirmed delta or omicron variant infections were included in the analysis.

This study was approved by the Institutional Review Board of Yonsei University Health System Clinical Trial Centre, and the study protocol adhered to the Declaration of Helsinki guidelines. Written informed consent was obtained from all the participants. (Approval number: 9-2021-0156, approved on 18^th^ November 2021)

### Data collection

We collected data on initial symptoms, reinfection, diagnosis date, initial PCR cycle threshold (Ct) value, COVID-19 vaccination history, and household contacts from the COVID-19 investigation report provided by the epidemiological investigator. The patients’ baseline characteristics and clinical course were recorded during hospitalisation. The Charlson comorbidity index (CCI) was used to categorise comorbidities in the patients[9]. Immunocompromised conditions were determined according to the severe immunosuppression definition by the UK Health Security Agency[10]. Vaccination status was divided into three groups—unvaccinated, vaccinated, and booster-vaccinated. The unvaccinated group included patients who had never been vaccinated, those who received one dose within the past three weeks, and those who were partially vaccinated (received just one dose or received two doses within the past two weeks). Vaccinated group included patients who received a vaccination ≥2 weeks ago; a single dose of Janssen Ad26.COV2.S vaccine was considered vaccinated if it had been more than 2 weeks after administration. Of the vaccinated patients, individuals were classified into the booster-vaccinated group if they received a booster shot before 2 weeks of enrolment.

### Procedures

Serum sample preparation, anti-SARS-CoV-2 antibody assays, RNA extraction, and PCR for SARS-CoV-2 detection and variant typing are described in appendix p 5-6.

### Outcomes

The study’s primary outcomes were to compare patients with delta and omicrappon variant infections and to determine the effect of vaccine-induced antibodies on the clinical courses of breakthrough infection caused by delta and omicron variants. The secondary outcome was to determine the viral dynamics according to the antibody level.

### Statistical analysis

Only patients who were tested for antibody levels within 7 days of symptom onset or diagnosis, whichever was earlier, were included in the analyses using antibody titres. The 7-day period was designated to minimise the effect of current infection on antibody levels elicited by vaccination[11]. Furthermore, we performed sensitivity analyses using 3-day and 5-day thresholds. We investigated viral dynamics using Ct values obtained from the PCR test. Viral dynamic analysis was conducted only on data from patients who had undergone the PCR test– 5–7 days after the initial diagnosis.

Continuous variables were analysed using descriptive methods depending on their distribution and tested using the Shapiro–Wilk test. Variables with a normal distribution were described as means and standard deviations, and independent two-sample t-tests were performed. Non-normal variables were expressed as medians and interquartile range (IQR). Categorical variables were described as frequencies and percentages. Chi-square or Fisher’s exact tests were performed depending on the number of expected events. Multivariable logistic regression analysis was performed to determine the effect of antibody levels on the clinical course of breakthrough infections. We selected fever (≥37.5 °C), hypoxia (≤94% oxygen saturation), pneumonia, C-reactive protein (CRP) elevation (>8 mg/L), and lymphopenia (<1,100 cells/μL) as variables representing the clinical course. Relevant variables with a significance level of less than 0.1 through univariate logistic regression analysis or with a clinical significance were included in the multivariable model. The relationship between antibody levels and Ct values was confirmed using a generalised linear model. Statistical analyses were performed using SAS (version 9.4; SAS Institute) and R (version 4.1.1; R Foundation for Statistical Computing).

## Results

A total of 187 patients were admitted to Yongin Severance Hospital between 11 December 2021 and 10 February 2022. Of these, 172 patients underwent tests for SARS-CoV-2 variant type assay. Results of the assay revealed that 79 patients were infected with the delta variant, 82 with the omicron variant, and seven with the undetermined type. There were no cases of re-infection. According to the inclusion criteria for analyses using antibody levels, 106 patients who underwent antibody testing within a defined period were selected from 111 vaccinated patients: 39 with delta variant infections and 67 with omicron variant infections (Figure 1A). Antibody levels were tested at a median of 4 days (IQR 2–6) of symptom onset or diagnosis, and variant-type assays were performed at 4 days (IQR 3–5) after the initial diagnosis (Figure 1B).

**Figure 1.**
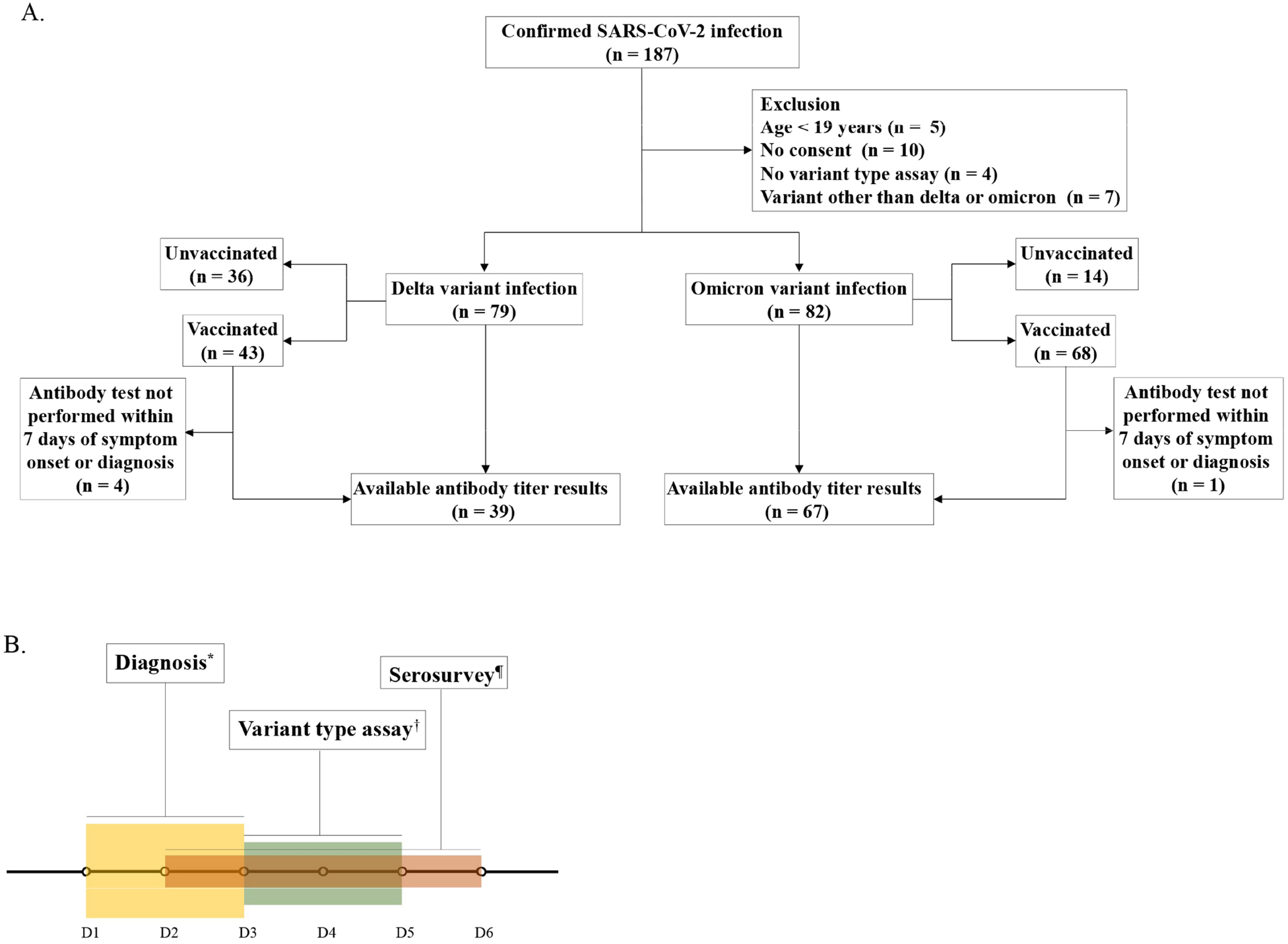
Study flow diagram. **A. Study flow of enrolment** **B. Severe acute respiratory syndrome coronavirus 2 diagnosis, antibody to spike protein test, variant type assay** *Patients were diagnosed at a median of 2 days (interquartile range [IQR] 1–3) after symptom onset. ¶Antibody tests were performed at a median of 4 days (IQR 2–6) after symptom onset or diagnosis, whichever was earlier. † Variant-type assays were conducted at a median of 4 days (IQR 3–5) after diagnosis.

Of the 161 patients who had the delta or omicron variant infection, 85 (53 %) were women, and the mean age was 54.5 (±18.9) years. Patients with omicron variant infection were younger than those with delta variant infection (p=0.017). The proportion of female patients, distribution of body mass index, frequency of immunocompromised status, or CCI ≥3 were similar among patients with delta and omicron variant infections. Although patients with delta variant infection were more frequently asymptomatic at the time of diagnosis (p=0.01), those with an omicron variant infection had lower occurrence rates of pneumonia (p<0.001) and hypoxia (p<0.001) during hospitalisation. Once patients experienced hypoxia, the duration of hypoxia was longer in patients with delta infections than in those with omicron variant infections (p=0.002). Compared with patients with omicron variant infection, those with delta variant infection had higher levels of CRP (p<0.001) and interleukin 6 (IL-6) (p=0.021) and were more likely to receive remdesivir (p=0.029), dexamethasone (p=0.002), and antimicrobial agents (p=0.001) (Table 1).

**Table 1.** Clinical characteristics of patients with SARS-CoV-2 infection according to variant type.

| Variable | Delta variant<br>(n = 79) | Omicron variant<br>(n = 82) | *p*-value |
| --- | --- | --- | --- |
| Demographic data |  |  |  |
| Age (years) | 58.1 ± 15.7 | 51.0 ± 21.1 | 0.017 |
| Female, n (%) | 40 (51.3%) | 45 (54.9%) | 0.590 |
| BMI (kg/m<sup>2</sup>) | 23.6 ± 3.1 | 23.7 ± 4.5 | 0.818 |
| Immunocompromised, n (%) | 11 (13.9%) | 12 (14.6%) | 0.898 |
| CCI ≥ 3, n (%) | 39 (49.4%) | 28 (34.1%) | 0.050 |
| Asymptomatic at diagnosis, n (%) | 14 (17.7%) | 4 (4.9%) | 0.010 |
| Laboratory data (worst results during hospitalisation) |  |  |  |
| CRP (mg/L) | 47.6 ± 65.8 | 15.6 ± 23.1 | <0.001 |
| Lymphocyte (10<sup>3</sup> cells/uL) | 1.1 ± 0.6 | 1.4 ± 0.6 | 0.001 |
| IL-6 (pg/mL) | 33.8 ± 73.7 | 9.8 ± 14.6 | 0.021 |
| D-dimer (mcgFEU/mL) | 0.8 ± 1.4 | 0.6 ± 1.1 | 0.276 |
| Clinical courses during hospitalisation |  |  |  |
| Fever (BT ≥ 37.5 °C), n (%) | 57 (72.2%) | 50 (61.7%) | 0.161 |
| Time to defervescence (days) | 4.6 ± 2.9 | 3.6 ± 2.3 | 0.065 |
| Pneumonia<sup>†</sup>, n (%) | 44 (55.7%) | 12 (14.6%) | <0.001 |
| Hypoxia (SpO<sub>2</sub> < 94 %), n (%) | 20 (25.3%) | 4 (4.9%) | <0.001 |
| Duration of oxygenation (days) | 6.6 ± 2.8 | 4.0 ± 0.0 | 0.002 |
| Treatments |  |  |  |
| High flow, n (%) | 3 (3.8%) | 1 (1.2%) | 0.361<sup>a</sup> |
| Regdanvimab, n (%) | 8 (10.1%) | 3 (3.7%) | 0.104 |
| Remdesivir, n (%) | 19 (24.1%) | 9 (11.0%) | 0.028 |
| Dexamethasone, n (%) | 28 (35.4%) | 12 (14.6%) | 0.002 |
| Antimicrobial agents, n (%) | 33 (41.8%) | 15 (18.3%) | 0.001 |

Data are expressed as mean ± standard deviation or number with percentages. Abbreviations: BMI, body mass index; CCI, Charlson comorbidity index; CRP, C-reactive protein; IL, interleukin; BT, body temperature; SpO<sub>2</sub>, percutaneous oxygen saturation; SARS-CoV-2, severe acute respiratory syndrome coronavirus 2
<sup>a</sup>*p*-value was calculated using Fisher's exact test.
<sup>†</sup>Presence of pneumonia was determined using chest radiography.

There were 111 vaccinated patients with delta variant (n=43) and omicron variant (n=68) infections. The comparison of delta and omicron variant infections in these patients was similar to that in 161 patients whose vaccination status was not classified (Supplementary Table 3). Unvaccinated patients (n=50) more commonly experienced fever, pneumonia, or hypoxia during hospitalisation than vaccinated patients did. Vaccinated patients had a shorter time to defervescence than unvaccinated ones. Laboratory results did not differ between the vaccinated and unvaccinated patients, except for lymphocyte counts. Vaccinated patients were less likely to be treated with drugs for COVID-19 than were unvaccinated patients (Supplementary Table 4).

When we classified vaccinated patients with delta and omicron variant infection into booster-vaccinated (n=47) and booster-unvaccinated (n=64) groups, as expected from a recent national vaccination program recommending a booster shot, the time since the last vaccination to confirm SARS-CoV-2 infection was shorter in the booster-vaccinated group than in the booster-unvaccinated group. Symptoms, signs, laboratory results, and treatment drugs, except for dexamethasone, did not differ between groups (Supplementary Table 5). Comparisons of vaccinated and unvaccinated patients or booster-unvaccinated and booster-vaccinated patients in each variant type are shown in Supplementary Tables 6–9.

Data from 106 patients whose serum samples were collected within seven days of symptom onset or diagnosis were used for analyses using antibody titres (Figure 1B). The geometric mean antibody titres in patients who experienced fever, hypoxia, pneumonia, CRP elevation, and lymphopenia during hospitalisation were 1,201.5 U/mL, 98.8 U/mL, 774.1 U/mL, 1,335.1 U/mL, and 1,032.2 U/mL, respectively, which were lower than those in patients who did not (Supplemental table 10 and Figure 2). Similar results were observed when the selected patients were divided into delta and omicron variant infection groups. Sensitivity analyses using 3-day and 5-day thresholds for time since symptom onset or diagnosis to antibody test did not differ from the main analyses (Supplementary Figures 2 and 3).

**Figure 2.**
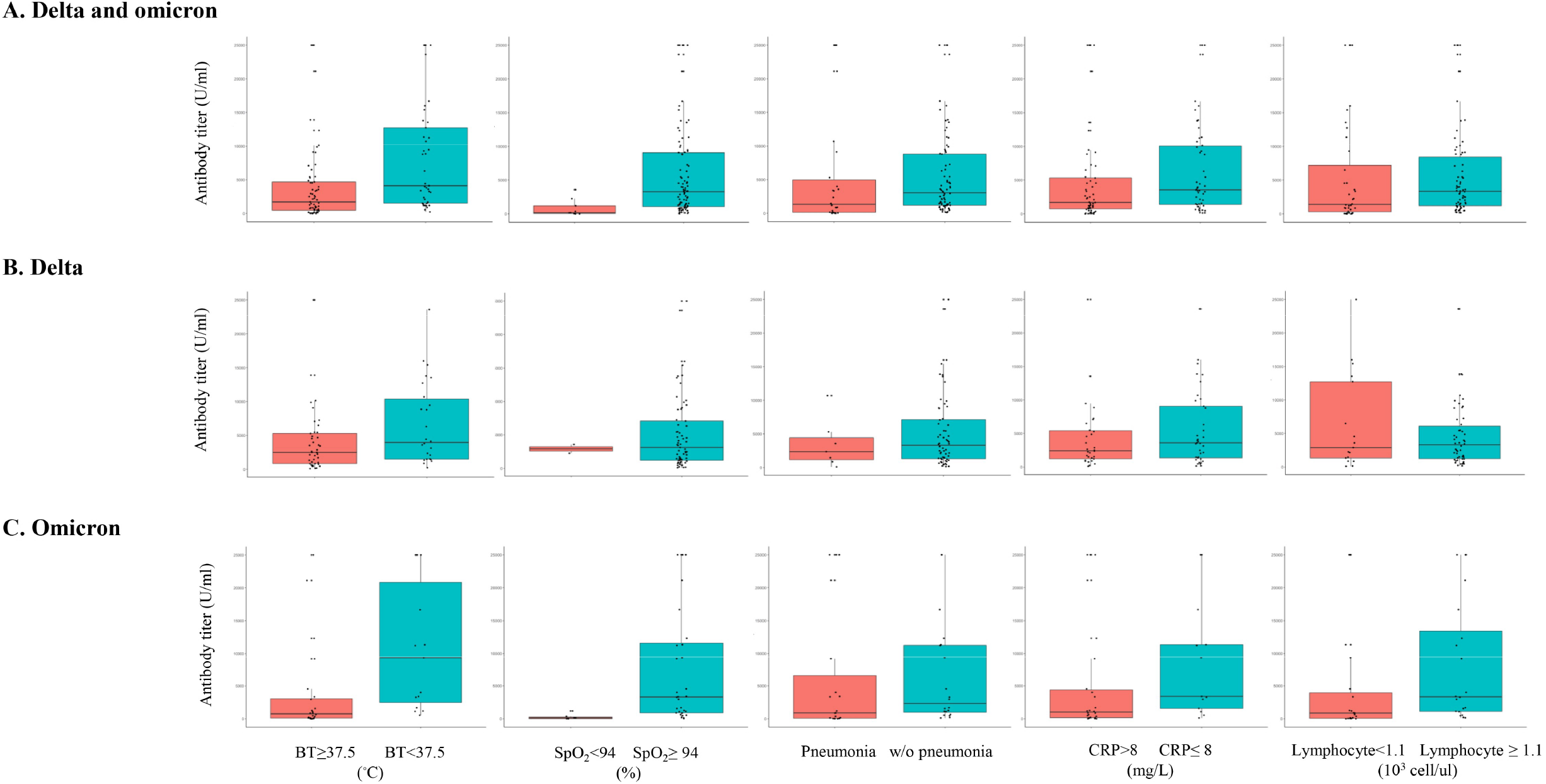
Comparison of antibody levels between vaccinated patients with or without specific signs during hospitalisation. This analysis included 106 patients with delta and omicron variant infections whose serum samples were collected within 7 days of symptom onset or diagnosis. Antibody levels are described as box plots of medians with interquartile ranges. BT, body temperature; SpO_2_, percutaneous oxygen saturation; w/o, without; CRP, C-reactive protein

The multivariable model showed that an increase in antibody levels in vaccinated patients with delta or omicron variant infection was independently associated with a decrease in the occurrence of fever (adjusted odds ratio [aOR], 0.231; 95% confidence interval [CI], 0.105– 0.511), hypoxia (aOR, 0.229; 95% CI, 0.075–0.703), CRP elevation (aOR, 0.524; 95% CI, 0.293–0.0.938), and lymphopenia (aOR, 0.568; 95% CI, 0.33–0.0.976) during hospitalisation (Table 2). Although the occurrence of pneumonia was associated with antibody levels in the unadjusted analysis, statistical significance was not observed in the adjusted analysis (aOR, 0.526; 95% CI, 0.249–1.112).

**Table 2.**
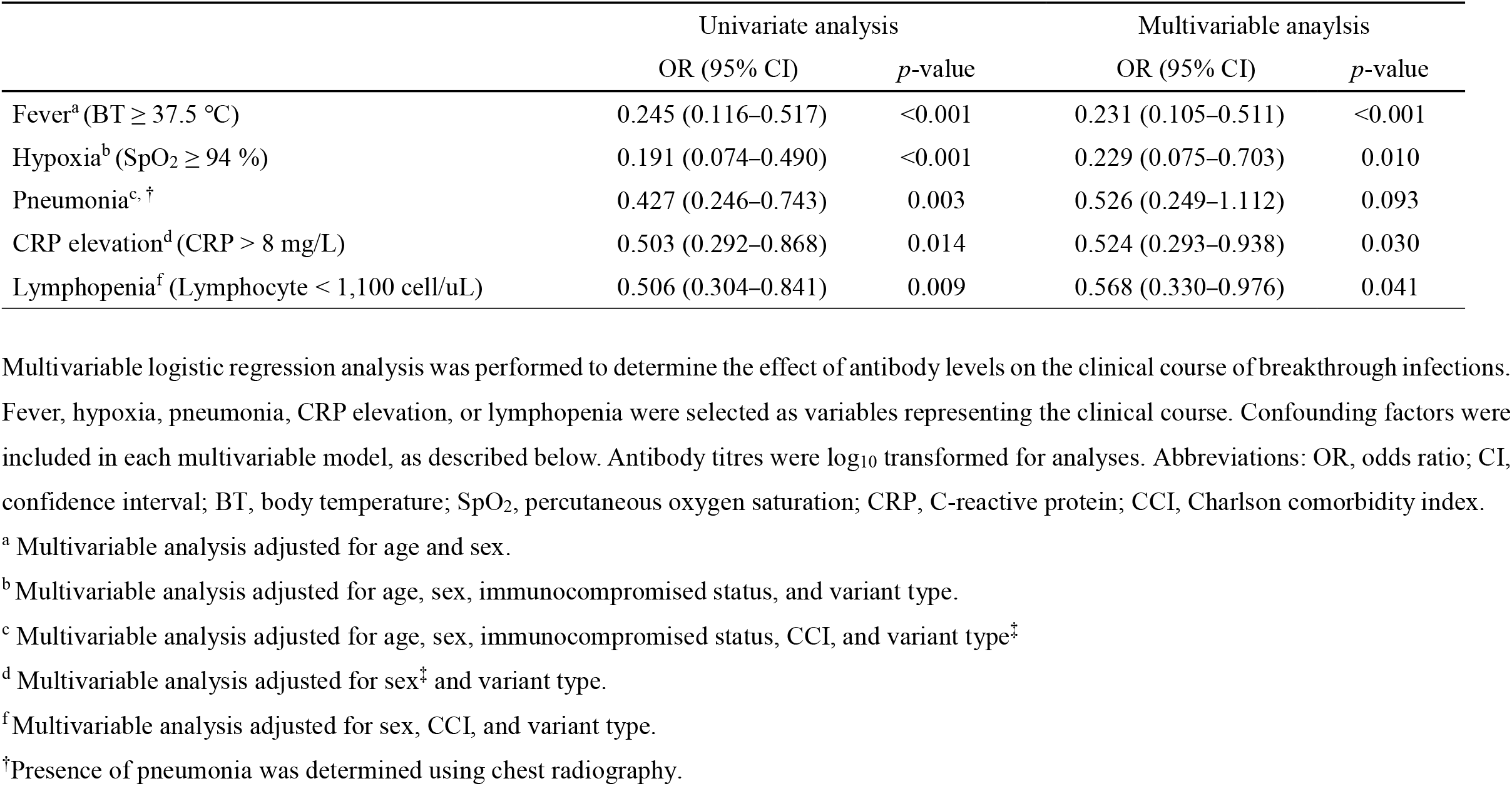
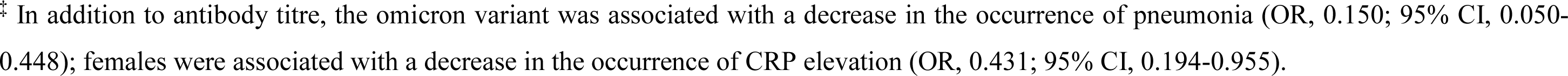
Association of antibody titres and variables with clinical courses during hospitalisation in vaccinated patients with breakthrough infections caused by delta and omicron variants.

|  | Univariate analysis |  | Multivariable analysis |  |
| --- | --- | --- | --- | --- |
|  | OR (95% CI) | *p*-value | OR (95% CI) | *p*-value |
| Fever<sup>a</sup> (BT $\geq 37.5$ °C) | 0.245 (0.116–0.517) | <0.001 | 0.231 (0.105–0.511) | <0.001 |
| Hypoxia<sup>b</sup> (SpO<sub>2</sub> $\geq 94$ %) | 0.191 (0.074–0.490) | <0.001 | 0.229 (0.075–0.703) | 0.010 |
| Pneumonia<sup>c, †</sup> | 0.427 (0.246–0.743) | 0.003 | 0.526 (0.249–1.112) | 0.093 |
| CRP elevation<sup>d</sup> (CRP > 8 mg/L) | 0.503 (0.292–0.868) | 0.014 | 0.524 (0.293–0.938) | 0.030 |
| Lymphopenia<sup>f</sup> (Lymphocyte < 1,100 cell/uL) | 0.506 (0.304–0.841) | 0.009 | 0.568 (0.330–0.976) | 0.041 |

Multivariable logistic regression analysis was performed to determine the effect of antibody levels on the clinical course of breakthrough infections. Fever, hypoxia, pneumonia, CRP elevation, or lymphopenia were selected as variables representing the clinical course. Confounding factors were included in each multivariable model, as described below. Antibody titres were log<sub>10</sub> transformed for analyses. Abbreviations: OR, odds ratio; CI, confidence interval; BT, body temperature; SpO<sub>2</sub>, percutaneous oxygen saturation; CRP, C-reactive protein; CCI, Charlson comorbidity index.
<sup>a</sup> Multivariable analysis adjusted for age and sex.
<sup>b</sup> Multivariable analysis adjusted for age, sex, immunocompromised status, and variant type.
<sup>c</sup> Multivariable analysis adjusted for age, sex, immunocompromised status, CCI, and variant type<sup>‡</sup>
<sup>d</sup> Multivariable analysis adjusted for sex<sup>‡</sup> and variant type.
<sup>f</sup> Multivariable analysis adjusted for sex, CCI, and variant type.
<sup>†</sup> Presence of pneumonia was determined using chest radiography.
<sup>‡</sup> In addition to antibody titre, the omicron variant was associated with a decrease in the occurrence of pneumonia (OR, 0.150; 95% CI, 0.050-0.448); females were associated with a decrease in the occurrence of CRP elevation (OR, 0.431; 95% CI, 0.194-0.955).

Therefore, we investigated the association between antibody levels and viral dynamics. Among 106 patients included in the analyses using antibody levels, 33 patients who had Ct values from the PCR tests 5–7 days after the initial diagnosis were selected. Patients with higher antibody levels had higher Ct values at 5–7 days after the initial diagnosis, indicating lower viral loads than those with lower antibody levels (p=0.022) (Figure 3). We also evaluated the association between antibody levels and Ct values at the initial diagnosis. Although several different commercial PCR kits were used at initial diagnosis because the patients were tested in different institutions at the time of diagnosis, we found no difference in Ct values at the time of diagnosis according to the antibody level (data not shown).

**Figure 3.**
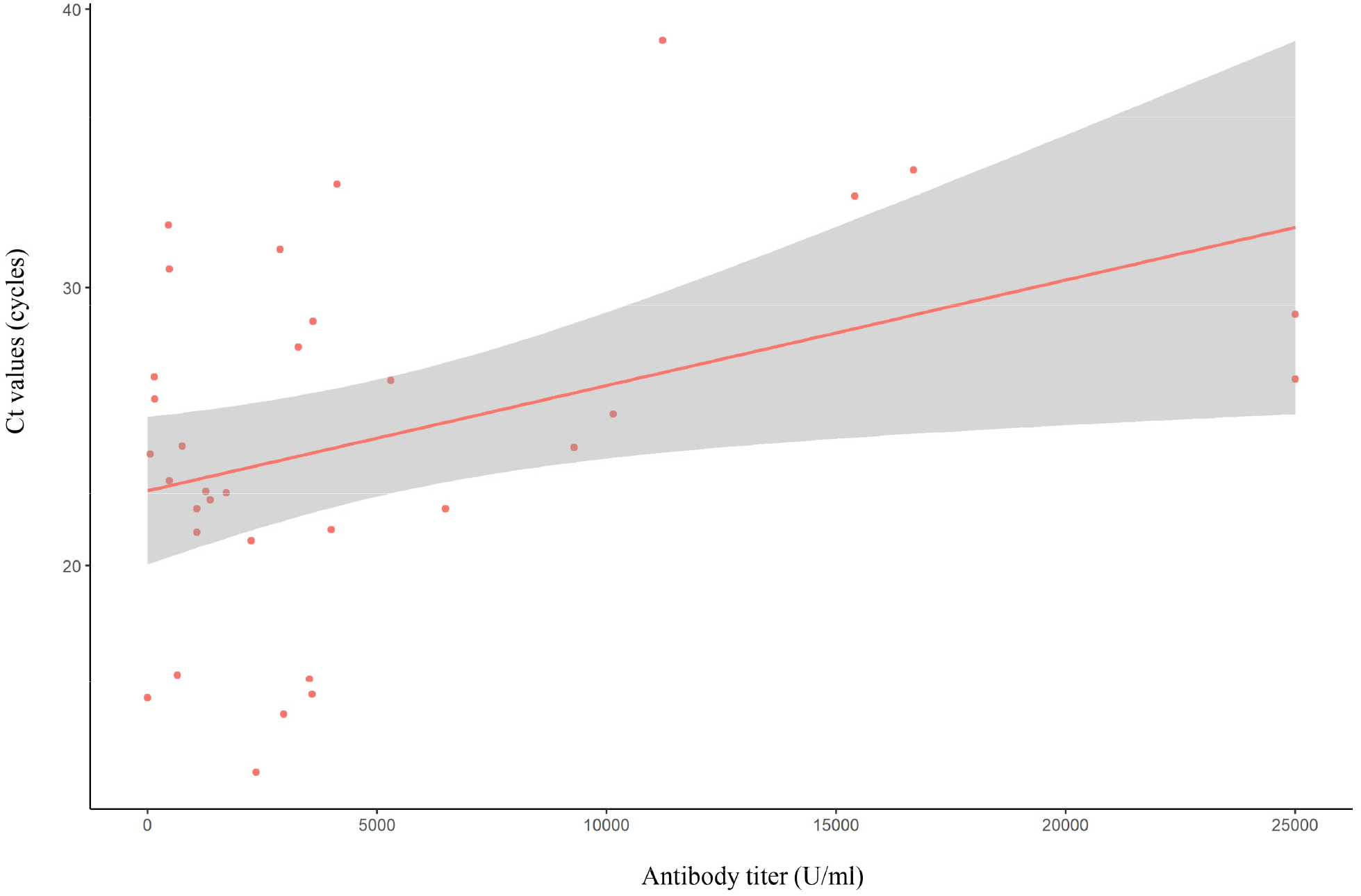
Association of antibody titres and Ct values. Data from 33 patients, with Ct values measured 5–7 days after diagnosis, showed a positive correlation between antibody levels and Ct values (slope: 0.0004, p=0.022) Abbreviations: Ct, cycle threshold; PCR, polymerase chain reaction

## Discussion

We identified that the vaccine was still effective against SARS-CoV-2 infection, including delta and omicron variants. Furthermore, our data showed that vaccine-induced antibody titres were independently associated with the occurrence of specific signs, as indicators of an unfavourable clinical course, in patients with SARS-CoV-2 breakthrough infection, regardless of virus type or booster vaccination status. Our study provides important information for taking measures to minimise the damage caused by the current pandemic.

Vaccines against SARS-CoV-2 are effective in the prevention of adverse outcomes in patients with infections, such as hospitalisation and death[12,13]. Neutralising antibody responses are highly predictive of protection in vaccinated individuals[14]. However, identifying neutralising antibodies is not feasible for practical use in clinical settings because it is a labour-intensive and time-consuming task. Instead, monitoring antibody levels to spike proteins using commercial kits is more convenient and logistically feasible. Our study provides evidence that antibody level is an important predictor of the clinical course in patients with breakthrough infection by showing a close relationship between low antibody levels and an unfavourable disease course such as fever, hypoxia, pneumonia, CRP elevation, and lymphopenia. Our data fill the gap of missing data on the impact of variable antibody responses to vaccination on the disease course of COVID-19.

Booster vaccination increases humoral immunity to prevent hospitalisation or severe disease progression in patients with omicron variant infection as well as delta-variant infection[15,16]. We also observed higher antibody levels in vaccine-completed individuals with primary vaccination than in booster-vaccinated individuals (data not shown). However, booster vaccination was not associated with clinical protection in patients with breakthrough infections caused by the delta and omicron variants in this study. We postulate that this finding is attributable to the fact that the indicator of clinical protection used in this study could be sufficiently prevented by the level of antibodies that was mounted by primary vaccination. This study could not evaluate the occurrence of severe outcomes (e.g. mechanical ventilation or mortality) as indicators of clinical protection because of the specific baseline conditions of the included patients. Therefore, the effect of antibodies according to different titres on these outcomes should be evaluated in future studies involving other populations.

Previous studies have reported that the omicron variant has weakened virulence, which in turn results in reduced severity even if infection occurs [17,18]. We also found similar findings in omicron variant infections compared with delta variant infections (Table 1). However, in breakthrough infections caused by delta and omicron variants, the variant type was not associated with the occurrence of specific signs observed in this study, except for pneumonia (Table 2). We assume that our findings are owing to the reduced effectiveness of vaccines against omicron variants. Indeed, the omicron variant has multiple mutations that enable evasion of the vaccine-induced immune system [19,20]. A decrease in vaccine-induced immunity against symptomatic omicron variant infection may offset the protective effect of the impaired virulence of the omicron variant. Our postulation is required to be corroborated by further studies that have an extended period of observation to determine the effect of variant type on the clinical course.

We found differences in the viral kinetics among vaccinated patients with a variety of antibody levels. The viral load decline rate was positively correlated with antibody levels. In other words, more rapid viral clearance was observed in patients with higher antibody levels than in those with lower antibody levels. These findings are in line with those of previous studies, which suggest that the host immune system determines viral clearance [21,22]. Although we did not evaluate the association of clinical courses and viral kinetics in this study owing to the limited number of participants who underwent PCR test 5–7 days after initial diagnosis, it is possible that rapid viral clearance contributes to the occurrence of specific signs observed in this study among patients with breakthrough infection. This hypothesis should be tested in future studies.

Our study had several limitations. First, we could not exclude the effects of previous or current infections on antibody levels. However, none of the participants had a previous infection, although it relied on self-reporting. Furthermore, our findings were robust in a sensitivity analysis involving patients who tested antibody levels within 3 or 5 days of symptom onset or diagnosis. Second, since our study hospital has specific criteria for admission, which allowed only patients with mild to moderate severity to be admitted, we could not investigate the effects of antibody levels on outcomes such as moving to the severe phase (n=2) or death (n=0). Therefore, further studies involving a larger number of patients with a wide range of disease severity are warranted. Third, the diagnosis of pneumonia in this study was based on chest radiographic findings. Chest radiography is limited in its ability to detect subtle pneumonia. Furthermore, we did not routinely perform follow-up chest radiography if the initial test did not report abnormal findings and if the patient’s condition did not change. Therefore, the results associated with pneumonia should be cautiously interpreted. Fourth, we did not consider the type of vaccine (e.g. adenoviral vector-based, mRNA-based) when analysing antibody levels. Most previous studies have demonstrated the effectiveness of vaccines based on homologous vaccination data. However, the real-world population receives heterologous vaccines from different platforms for several reasons, such as the disruption of vaccine supply and vaccine-associated adverse events. Moreover, booster immunisation programs have increased the possibility of receiving different types of vaccines. In this regard, we did not classify the participants according to the type of vaccine but only vaccination status to reflect real-world population data.

In conclusion, this study showed the significance of vaccine-induced antibody levels as a predictor of the clinical course of COVID-19 in patients with breakthrough infections caused by delta and omicron variants. Moreover, in our cohort, enhanced viral clearance was observed in vaccinated patients with higher antibody levels than in those with lower antibody levels. We highlight the need for concentrated efforts to monitor patients with SARS-CoV-2 infection who have conditions related to poor antibody response to vaccination (e.g. immunocompromised status) or waning immunity after vaccination (e.g. adenoviral vector-based vaccine) since they are more likely to undergo an unfavourable disease course.

## Data Availability

The dataset supporting the conclusions of this article is included within Supplemental data

## Acknowledgments

We would like to thank all of the nursing and laboratory staff as well as the physicians who supported this project. Finally, we give credit to all of the patients who took part in this study.

## Availability of data and materials

The dataset supporting the conclusions of this article is included within Supplemental data (Raw data used in analysis).

