## Supplemental tables and figures for "Vaccine-induced antibody level predicts the clinical course of breakthrough infection of COVID-19 caused by delta and omicron variants: a prospective observational cohort study"

**Supplementary appendix**

This appendix has been provided by the authors to supplement readers with additional information about this work.

### 1. Clinical symptoms and risk factors for which hospitalization is considered in confirmed COVID-19 patients

Patients where the following applies are asked to be hospitalized for monitoring and treatment. In the early stages of COVID-19, Korea mandated that all confirmed patients should be hospitalised or enter a treatment centre. Since October 2021, the concept of home treatment has been actively used to support the efficient use of available hospital beds. Since November 2021, home treatment has been widely preferred; only cases that fall under the following indications are hospitalised.

**Supplementary Table 1. Clinical symptoms and risk factors for which hospitalisation is considered in confirmed COVID-19 patients**

- A person aged 70 and older who is not vaccinated
- Unconsciousness after the appearance of COVID-19 symptoms
- Difficulty in breathing (in daily life)
- Fever higher than 38 °C that is not controlled by antipyretic drugs
- Diabetes that is not controlled by drug use
- A patient who needs dialysis
- Chronic lung disease, asthma, heart failure, or coronary artery disease treated with drugs after diagnosis
- A patient receiving chemotherapy or immunosuppressive drugs
- A patient with mental illness uncontrolled by drugs
- A person who spends more than 50% of the daytime in bed
- A person with severe obesity (BMI above 30)
- A pregnant woman with symptoms (abdominal pain, labour pain, vaginal bleeding, etc.)
- High-risk and severely ill children
  - ■ Difficulty in breathing, dyspnoea, cyanosis
  - ■ Poor oral intake and dehydration
  - ■ Children diagnosed with chronic lung disease/heart disease/metabolic disease/ immune disorder and receiving immunosuppressants
  - ■ Children with respiratory function disorder, secretion discharge, or high risk of aspiration

**2. Severity of COVID-19**

The most important factor in determining whether hospitalisation is necessary for a confirmed case is the severity of symptoms. The patients were assigned to appropriate hospitals according to the severity of COVID-19.

**Supplementary Table 2. Disease severity classification for COVID-19**

| Level | Definition | Severity |
|---|---|---|
| 1 | No limit of activity | Mild or less |
| 2 | Limited activity but no $O_2$ needed | |
| 3 | $O_2$ with nasal prong | Moderate |
| 4 | $O_2$ with facial mask | |
| 5 | Non-invasive ventilation/high-flow $O_2$ | Severe |
| 6 | Invasive ventilation | |
| 7 | Multi-organ failure/ECMO/CRRT | |
| 8 | Death | Death |

**3. Percentage of omicron variant infection cases in South Korea**

**Supplementary Figure 1. Percentage of omicron variant infection cases in South Korea**

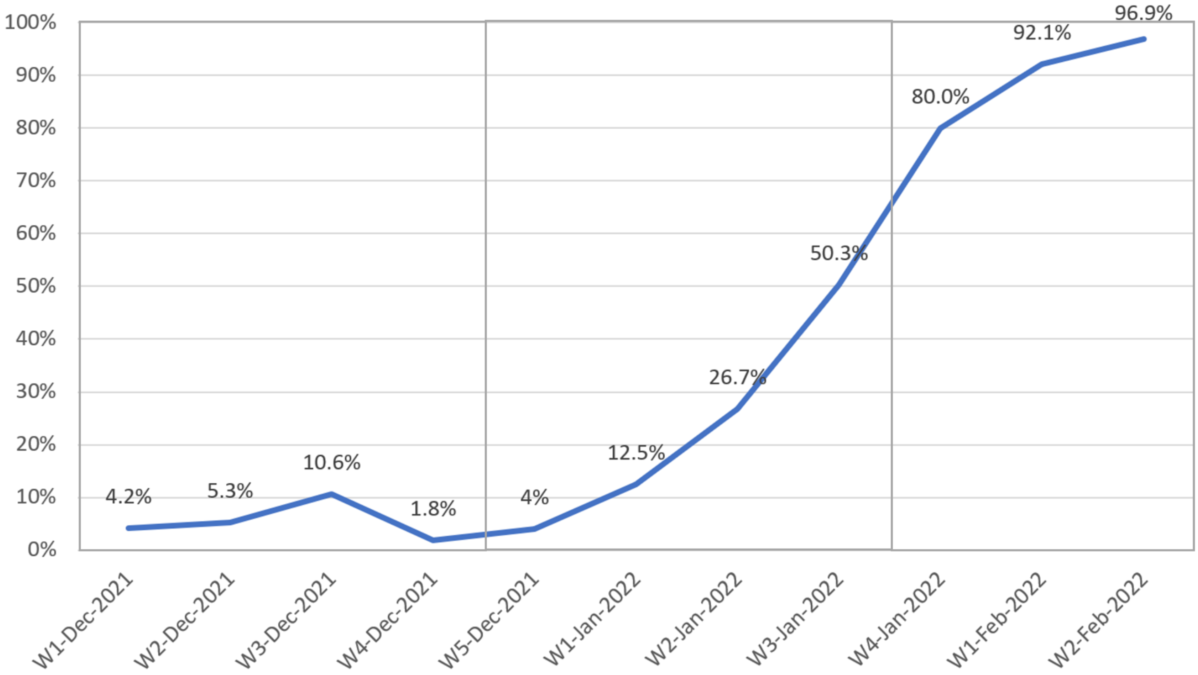


In contrast to other countries, in which the omicron variant became the dominant strain within 4–6 weeks, it took about nine weeks to become the dominant strain since the first notification of the omicron variant on 24 November 2021 due to quarantine and isolation guidelines in South Korea. (Source: Korea Disease Control and Prevention Agency press release, 14 January 2022)

### 4. Laboratory procedures

#### 4.1. Sample preparation

Serum samples for antibody measurement were collected on the day of admission and stored at -20 °C until analysis was performed. Frozen samples were thawed before antibody determination.

#### 4.2. Anti-SARS-CoV-2 assays

Antibodies against the S protein, the target of vaccines, were analysed using the Elecsys AntiSARS-CoV-2 S assay (Roche Diagnostics, Mannheim, Germany), an electrochemiluminescence immunoassay that detects antibodies, including IgG, to the receptor-binding domain (RBD) of the SARS-CoV-2 spike protein on the Cobas e801 module (Roche Diagnostics, Mannheim, Germany), with a measuring range from 0.4 U/mL to 250 U/mL (up to 25,000 U/mL with onboard 1:100 dilution). Values ≥0.8 U/mL were considered positive. All samples were processed according to the manufacturer's instructions with specified controls and calibrators by trained laboratory staff.

#### 4.3. RNA Extraction and SARS-CoV-2 and Variants Detection

The MagNA Pure 96 System (Roche Diagnostics, Mannheim, Germany) was used to extract RNA from nasopharyngeal swabs in viral transport media, according to the manufacturer's instructions. The extracted RNA was then subjected to the Allplex SARS-CoV-2 Assay, which targets four genes in a single tube (E, N, RdRp, and S genes) to detect SARS-CoV-2 infection, according to the manufacturer's instructions. PCR amplification was performed using the CFX96 real-time PCR detection system (Bio-Rad Laboratories, Hercules, CA, USA).

For SARS-CoV-2 variant detection, Novaplex SARS-CoV-2 Variants I, II, IV, and VII assays

(Seegene, Seoul, South Korea) were used in combination, according to the manufacturer's instructions. Briefly, 5 μL of the nucleic acid extract was mixed with 5 μL of primers and probes, 5 μL of buffer, and 5 μL of the enzyme contained in each kit. Plates were run on CFX96 with the following protocol: a reverse transcription step was initially performed at 50 °C for 20 min, then a denaturation step 95 °C for 15 min, initial cycling for 3 steps at 95 °C for 10 s, 60 °C for 40 s, and 72 °C for 20 s were performed. Finally, 42 rapid cycles at 95 °C for 10 s, 60 °C for 15 s, and 72 °C for 10 s were executed and fluorescence reads were recorded at 60 °C and 72 °C. Results interpretation and data analysis were performed using Seegene viewer. Detectable target mutations for each kit are as follows: 69/70del, E484K, and N501Y for the Variants I; W152C K417N, K417T and L452R for the Variants II; K417N, L452R, and P681R for the Variants IV; 69/70del, E484A, and N501Y for the Variants VII Assay. The target and detectable variants for each assay are shown in the Supplementary Table.

### 5. Supplementary Tables and Figure legends

**Supplementary Table 3. Clinical characteristics of patients with SARS-CoV-2 breakthrough infection according to variant type**

| Variable | Delta variant ($n$ = 43) | Omicron variant ($n$ = 68) | $p$-value |
|---|---|---|---|
| Demographic data | | | |
| Age (years) | 60.7 ± 14.9 | 53.2 ± 21.2 | 0.032 |
| Female, n (%) | 18 (41.9%) | 37 (54.4%) | 0.197 |
| Body weight index (kg/m$^2$) | 24.3 ± 3.4 | 23.3 ± 3.8 | 0.143 |
| Immunocompromised, n (%) | 6 (14.0%) | 7 (10.3%) | 0.559 |
| Charlson comorbidity index ≥ 3, n (%) | 24 (55.8%) | 27 (39.7%) | 0.097 |
| Asymptomatic at diagnosis, n (%) | 9 (20.9%) | 2 (2.9%) | 0.003[a] |
| Time since the completion of vaccination (days) | 93 ± 59.4 | 85.6 ± 48.0 | 0.431 |
| Laboratory data (worst results during hospitalisation) | | | |
| CRP (mg/L) | 52.0 ± 79.4 | 13.9 ± 20.3 | 0.003 |
| Lymphocyte ($10^3$cells/uL) | 1.3 ± 0.7 | 1.41 ± 0.5 | 0.288 |
| IL-6 (pg/mL) | 44.9 ± 96.6 | 8.8 ± 12.6 | 0.047 |
| D-dimer (mcgFEU/mL) | 0.8 ± 1.6 | 0.6 ± 1.1 | 0.328 |
| Clinical courses during hospitalisation | | | |
| Fever (BT ≥ 37.5 ℃), n (%) | 26 (60.5%) | 41 (60.3%) | 0.986 |
| Time to defervescence (days) | 3.9 ± 2.2 | 3.3 ± 1.8 | 0.278 |

| | | | |
|---|---|---|---|
| Pneumonia[†], n (%) | 21 (48.8%) | 7 (10.3%) | <0.001 |
| Hypoxia ($SpO_2$ < 94 %), n (%) | 9 (20.9%) | 2 (2.9%) | 0.003[a] |
| Duration of oxygenation (days) | 7 ± 2.7 | 4.0 ± 0.0 | 0.374 |
| Treatments | | | |
| High flow, n (%) | 2 (4.7%) | 0 (0.0%) | 0.148[a] |
| Regdanvimab, n (%) | 3 (7.0%) | 2 (2.9%) | 0.374[a] |
| Remdesivir, n (%) | 9 (20.9%) | 5 (7.4%) | 0.036 |
| Dexamethasone, n (%) | 12 (27.9%) | 7 (10.3%) | 0.016 |
| Antimicrobial agents, n (%) | 16 (37.2%) | 10 (14.7%) | 0.006 |

Data are expressed as mean ± standard deviation or number with percentages. Abbreviations: BMI, body mass index; CCI, Charlson comorbidity index; CRP, C-reactive protein; IL, interleukin; BT, body temperature; $SpO_2$, percutaneous oxygen saturation; SARS-CoV-2, severe acute respiratory syndrome coronavirus 2

[a]*p*-value was calculated by Fisher's Exact Test.

[†]Presence of pneumonia was determined using chest radiography.

**Supplementary Table 4. Clinical characteristics of patients with delta and omicron variant infection according to vaccination status**

| Variable | Unvaccinated ($n$ = 50) | Vaccinated ($n$ = 111) | $p$-value |
|---|---|---|---|
| Demographic data | | | |
| Age (years) | 50.9 ± 17.7 | 56.1 ± 19.3 | 0.108 |
| Female, n (%) | 30 (60.0%) | 55 (49.6%) | 0.219 |
| Body weight index (kg/m$^2$) | 23.5 ± 4.4 | 23.7 ± 3.7 | 0.806 |
| Immunocompromised, n (%) | 10 (20.0%) | 13 (11.7%) | 0.164 |
| Charlson comorbidity index ≥ 3, n (%) | 16 (32.0%) | 51 (46.0%) | 0.097 |
| Asymptomatic at diagnosis, n (%) | 43 (86.0%) | 100 (90.1%) | 0.446 |
| Laboratory data (worst results during hospitalisation) | | | |
| CRP (mg/L) | 37.1 ± 42.7 | 28.6 ± 54.8 | 0.337 |
| Lymphocyte (10$^3$cells/uL) | 1.0 ± 0.6 | 1.4 ± 0.6 | 0.002 |
| IL-6 (pg/mL) | 17.6 ± 17.6 | 20.7 ± 58.3 | 0.644 |
| D-dimer (mcgFEU/mL) | 0.7 ± 1.0 | 0.6 ± 1.3 | 0.999 |
| Clinical courses during hospitalisation | | | |
| Fever (BT ≥ 37.5 ℃), n (%) | 41 (82.0%) | 67 (60.4%) | 0.007 |
| Time to defervescence (days) | 5.1 ± 3.3 | 3.5 ± 2.0 | 0.011 |
| Pneumonia[†], n (%) | 28 (56.0%) | 28 (25.2%) | <0.001 |
| Hypoxia ($SpO_2$ < 94 %), n (%) | 13 (26.0%) | 11 (9.9%) | 0.008 |
| Duration of oxygenation (days) | 6.3 ± 2.9 | 6.5 ± 2.7 | 0.862 |
| Treatments | | | |
| High flow, n (%) | 2 (4.0%) | 2 (1.8%) | 0.589[a] |
| Regdanvimab, n (%) | 6 (12.0%) | 5 (4.5%) | 0.098[a] |

| | | | |
|---|---|---|---|
| Remdesivir, n (%) | 14 (28.0%) | 14 (12.6%) | 0.017 |
| Dexamethasone, n (%) | 21 (42%) | 19 (17.1%) | <0.001 |
| Antimicrobial agents, n (%) | 22 (44%) | 26 23.4%) | 0.008 |

Patients were classified into two groups according to vaccination status, regardless of variant type. Data are expressed as mean ± standard deviation or number with percentages. Abbreviations: BMI, body mass index; CCI, Charlson comorbidity index; CRP, C-reactive protein; IL, interleukin; BT, body temperature; $SpO_2$, percutaneous oxygen saturation

[a] *p*-value was calculated by Fisher's Exact Test.

[†] Presence of pneumonia was determined using chest radiography.

**Supplementary Table 5. Clinical characteristics of patients with breakthrough infection caused by delta and omicron variant according to booster vaccination status**

| Variable | Booster-unvaccinated ($n$ = 64) | Booster-vaccinated ($n$ = 47) | $p$-value |
|---|---|---|---|
| Demographic data | | | |
| Age (years) | 49.1 ± 18.5 | 65.6 ± 16.1 | <0.001 |
| Female, n (%) | 29 (45.3%) | 26 (55.3%) | 0.298 |
| Body weight index ($kg/m^2$) | 23.5 ± 3.5 | 24.0 ± 3.9 | 0.453 |
| Immunocompromised, n (%) | 8 (12.5%) | 5 (10.7%) | 0.763 |
| Charlson comorbidity index ≥ 3, n (%) | 24 (37.5%) | 27 (57.5%) | 0.037 |
| Asymptomatic at diagnosis, n (%) | 6 (9.4%) | 5 (10.6%) | >0.999[a] |
| Time since the completion of vaccination (days) | 122.5 ± 39.1 | 42.9 ± 28.1 | <0.001 |
| Laboratory data (worst results during hospitalisation) | | | |
| CRP (mg/L) | 36.3 ± 67.2 | 18.2 ± 28.3 | 0.056 |
| Lymphocyte ($10^3$cells/uL) | 1.3 ± 0.6 | 1.5 ± 0.6 | 0.064 |
| IL-6 (pg/mL) | 27.9 ± 74.5 | 10.6 ± 16.1 | 0.100 |
| D-dimer (mcgFEU/mL) | 0.8 ± 1.6 | 0.5 ± 0.7 | 0.243 |
| Clinical courses during hospitalisation | | | |
| Fever (BT ≥ 37.5 ℃), n (%) | 41 (64.1%) | 26 (55.3%) | 0.352 |
| Time to defervescence (days) | 3.8 ± 2.3 | 3.1 ± 1.3 | 0.123 |
| Pneumonia[†], n (%) | 19 (29.7%) | 9 (19.2%) | 0.207 |
| Hypoxia ($SpO_2$ < 94 %), n (%) | 8 (12.5%) | 3 (6.38%) | 0.350[a] |
| Duration of oxygenation (days) | 7.8 ± 2.5 | 4.0 ± 0.0 | 0.058 |
| Treatments | | | |
| High flow, n (%) | 2 (3.1%) | 0 (0.0%) | 0.507[a] |

| | | | |
|---|---|---|---|
| Regdanvimab, n (%) | 3 (4.7%) | 2 (4.3%) | >0.999[a] |
| Remdesivir, n (%) | 10 (15.6%) | 4 (8.5%) | 0.265 |
| Dexamethasone, n (%) | 15 (23.4%) | 4 (8.5%) | 0.039 |
| Antimicrobial agents, n (%) | 18 28.1%) | 8 (17.0%) | 0.172 |

Patients were classified into two groups according to booster vaccination status, regardless of the variant type. Data are expressed as mean ± standard deviation or number with percentages. Abbreviations: BMI, body mass index; CCI, Charlson comorbidity index; CRP, C-reactive protein; IL, interleukin; BT, body temperature; $SpO_2$, percutaneous oxygen saturation

[a]*p*-value was calculated by Fisher's Exact Test.

[†]Presence of pneumonia was determined using chest radiography.

**Supplementary Table 6. Clinical characteristics of patients with delta variant infection according to vaccination status**

| Variable | Unvaccinated ($n$ = 36) | Vaccinated ($n$ = 43) | $p$-value |
|---|---|---|---|
| Demographic data | | | |
| Age (years) | 55 ± 16.2 | 60.7 ± 14.9 | 0.111 |
| Female, n (%) | 22 (61.1%) | 18 (41.9%) | 0.088 |
| Body weight index ($kg/m^2$) | 22.6 ± 2.5 | 24.3 ± 3.4 | 0.017 |
| Immunocompromised, n (%) | 5 (13.9%) | 6 (14.0%) | 0.993 |
| Charlson comorbidity index ≥ 3, n (%) | 15 (41.7%) | 24 (55.8%) | 0.210 |
| Asymptomatic at diagnosis, n (%) | 5 (13.9%) | 9 (20.9%) | 0.414 |
| Laboratory data (worst results during hospitalisation) | | | |
| CRP (mg/L) | 42.2 ± 45.2 | 52.0 ± 60.5 | 0.495 |
| Lymphocyte ($10^3$cells/uL) | 0.9 ± 0.4 | 1.3 ± 2.2 | 0.004 |
| IL-6 (pg/mL) | 19.4 ± 14.9 | 44.9 ± 96.6 | 0.157 |
| D-dimer (mcgFEU/mL) | 0.7 ± 1.2 | 0.8 ± 1.6 | 0.691 |
| Clinical courses during hospitalisation | | | |
| Fever (BT ≥ 37.5 °C), n (%) | 31 (86.1%) | 26 (60.5%) | 0.011 |
| Time to defervescence (days) | 5.1 ± 3.2 | 3.9 ± 2.2 | 0.135 |
| Pneumonia[†], n (%) | 23 (63.9%) | 21 (48.8%) | 0.180 |
| Hypoxia ($SpO_2$ < 94 %), n (%) | 11 (30.6%) | 9 (20.9%) | 0.327 |
| Duration of oxygenation (days) | 6.5 ± 2.9 | 7.0 ± 2.7 | 0.729 |
| Treatments | | | |
| High flow, n (%) | 1 (2.8%) | 2 (4.7%) | >0.999[a] |
| Regdanvimab, n (%) | 5 (13.9%) | 3 (7.0%) | 0.458[a] |

| | | | |
|---|---|---|---|
| Remdesivir, n (%) | 10 (27.8%) | 9 (20.9%) | 0.478 |
| Dexamethasone, n (%) | 16 (44.4%) | 12 (27.9%) | 0.126 |
| Antimicrobial agents, n (%) | 17 (47.2%) | 16 (37.2%) | 0.369 |

Data are expressed as mean ± standard deviation or number with percentages. Abbreviations: BMI, body mass index; CCI, Charlson comorbidity index; CRP, C-reactive protein; IL, interleukin; BT, body temperature; $SpO_2$, percutaneous oxygen saturation

[a] *p*-value was calculated by Fisher's Exact Test.

[†] Presence of pneumonia was determined using chest radiography.

**Supplementary Table 7. Clinical characteristics of patients with breakthrough infection caused by delta variant according to booster vaccination status**

| Variable | Fully vaccinated ($n$ = 30) | Booster vaccinated ($n$ = 13) | $p$-value |
|---|---|---|---|
| Demographic data | | | |
| Age (years) | 58.4 ± 15.7 | 65.9 ± 11.7 | 0.135 |
| Female, n (%) | 10 (33.3%) | 8 (61.5%) | 0.085 |
| Body weight index ($kg/m^2$) | 24.3 ± 3.3 | 24.4 ± 3.9 | 0.960 |
| Immunocompromised, n (%) | 4 (13.3%) | 2 (15.4%) | >0.999 |
| Charlson comorbidity index ≥ 3, n (%) | 18 (60.0%) | 6 (46.2%) | 0.401 |
| Asymptomatic at diagnosis, n (%) | 6 (20.0%) | 3 (23.1%) | >0.999[a] |
| Time since the completion of vaccination (days) | 121.9 ± 47.3 | 28.9 ± 19.7 | <0.001 |
| Laboratory data (worst results during hospitalisation) | | | |
| CRP (mg/L) | 63.5 ± 90.0 | 25.6 ± 37.4 | 0.058 |
| Lymphocyte ($10^3$cells/uL) | 1.2 ± 0.7 | 1.4 ± 0.7 | 0.302 |
| IL-6 (pg/mL) | 57.6 ± 112.2 | 13.7 ± 20.0 | 0.090 |
| D-dimer (mcgFEU/mL) | 1.0 ± 1.9 | 0.6 ± 0.7 | 0.360 |
| Clinical courses during hospitalisation | | | |
| Fever (BT ≥ 37.5 °C), n (%) | 20 (66.7%) | 6 (46.2%) | 0.206 |
| Time to defervescence (days) | 4.1 ± 2.5 | 3.2 ± 1.2 | 0.380 |
| Pneumonia[†], n (%) | 17 (56.7%) | 4 (30.8%) | 0.119 |
| Hypoxia ($SpO_2$ < 94 %), n (%) | 8 (26.7%) | 1 (7.7%) | 0.237[a] |
| Duration of oxygenation (days) | 7.8 ± 2.5 | 4.0 ± 0.0 | 0.272 |
| Treatments | | | |
| High flow, n (%) | 2 (6.7%) | 0 (0.0%) | >0.999[a] |

| | | | |
|---|---|---|---|
| Regdanvimab, n (%) | 3 (10.0%) | 0 (0.0%) | 0.542[a] |
| Remdesivir, n (%) | 8 (26.7%) | 1 (7.7%) | 0.237[a] |
| Dexamethasone, n (%) | 11 (36.7%) | 1 (7.7%) | 0.070[a] |
| Antimicrobial agents, n (%) | 14 (46.7%) | 2 (15.4%) | 0.086[a] |

Patients were classified into two groups according to their booster vaccination status. Data are expressed as mean ± standard deviation or number with percentages. Abbreviations: BMI, body mass index; CCI, Charlson comorbidity index; CRP, C-reactive protein; IL, interleukin; BT, body temperature; $SpO_2$, percutaneous oxygen saturation

[a]*p*-value was calculated by Fisher's Exact Test.

[†]Presence of pneumonia was determined using chest radiography.

**Supplementary Table 8. Clinical characteristics of patients with omicron variant infection according to vaccination status**

| Variable | Unvaccinated ($n$ = 14) | Vaccinated ($n$ = 68) | $p$-value |
|---|---|---|---|
| Demographic data | | | |
| Age (years) | 40.4 ± 17.5 | 53.2 ± 21.2 | 0.037 |
| Female, n (%) | 8 (57.1%) | 37 (54.4%) | 0.852 |
| Body weight index (kg/m$^2$) | 25.8 ± 6.9 | 23.3 ± 3.8 | 0.209 |
| Immunocompromised, n (%) | 5 (35.7%) | 7 (10.3%) | 0.028[a] |
| Charlson comorbidity index ≥ 3, n (%) | 1 (7.1%) | 27 (39.7%) | 0.028[a] |
| Asymptomatic at diagnosis, n (%) | 2 (14.3%) | 2 (2.9%) | 0.133[a] |
| Laboratory data (worst results during hospitalisation) | | | |
| CRP (mg/L) | 23.8 ± 33.6 | 13.9 ± 20.3 | 0.302 |
| Lymphocyte (10$^3$cells/uL) | 1.4 ± 0.7 | 1.4 ± 0.5 | 0.952 |
| IL-6 (pg/mL) | 14.4 ± 22.0 | 8.8 ± 12.6 | 0.397 |
| D-dimer (mcgFEU/mL) | 0.6 ± 0.5 | 0.6 ± 1.1 | 0.964 |
| Clinical courses during hospitalisation | | | |
| Fever (BT ≥ 37.5 ℃), n (%) | 10 (71.4%) | 41 (60.3%) | 0.434 |
| Time to defervescence (days) | 5.1 ± 3.9 | 3.3 ± 1.8 | 0.266 |
| Pneumonia[†], n (%) | 5 (35.7%) | 7 (10.3%) | 0.028[a] |
| Hypoxia ($SpO_2$ < 94 %), n (%) | 2 (14.3%) | 2 (2.9%) | 0.133[a] |
| Duration of oxygenation (days) | 4.0 ± 0.0 | 4.0 ± 0.0 | NA |
| Treatments | | | |
| High flow, n (%) | 1 (7.1%) | 0 (0.0%) | 0.171[a] |
| Regdanvimab, n (%) | 1 (7.1%) | 2 (2.9%) | 0.434[a] |

| | | | |
|---|---|---|---|
| Remdesivir, n (%) | 4 (28.6%) | 5 (7.4%) | 0.042[a] |
| Dexamethasone, n (%) | 5 (35.7%) | 7 (10.3%) | 0.028[a] |
| Antimicrobial agents , n (%) | 5 (35.7%) | 10 (14.7%) | 0.121 |

Data are expressed as mean ± standard deviation or number with percentages. BMI, body mass index; CCI, Charlson comorbidity index; CRP, C-reactive protein; IL, interleukin; BT, body temperature; $SpO_2$, saturation of percutaneous oxygen; NA, not applicable

[a]*p*-value was calculated by Fisher's Exact Test.

[†]Presence of pneumonia was determined using chest radiography.

**Supplementary Table 9. Clinical characteristics of patients with breakthrough infection caused by omicron variant according to booster vaccination status**

| Variable | Fully vaccinated ($n$ = 34) | Booster vaccinated ($n$ = 34) | $p$-value |
|---|---|---|---|
| Demographic data | | | |
| Age (years) | 40.9 ± 17.1 | 65.5 ±17.7 | <0.001 |
| Female, n (%) | 19 (55.9%) | 18 (52.9%) | 0.808 |
| Body weight index (kg/m$^2$) | 22.7 ±3.6 | 23.8 ± 3.9 | 0.218 |
| Immunocompromised, n (%) | 4 (11.8%) | 3 (8.8%) | >0.999[a] |
| Charlson comorbidity index ≥ 3, n (%) | 6 (17.7%) | 21 (61.8%) | <0.001 |
| Asymptomatic at diagnosis, n (%) | 0 (0.0%) | 2 (5.9%) | 0.492[a] |
| Time since the completion of vaccination (days) | 123.0 ± 30.9 | 48.3 ± 29.3 | <0.001 |
| Laboratory data (worst results during hospitalisation) | | | |
| CRP (mg/L) | 12.4 ± 15.9 | 15.4 ± 24.1 | 0.553 |
| Lymphocyte (10$^3$cells/uL) | 1.3 ± 0.5 | 1.5 ± 0.5 | 0.185 |
| IL-6 (pg/mL) | 8.1 ± 10.2 | 9.6 ± 15.0 | 0.636 |
| D-dimer (mcgFEU/mL) | 0.6 ± 1.4 | 0.5 ± 0.7 | 0.582 |
| Clinical courses during hospitalisation | | | |
| Fever (BT ≥ 37.5 ℃), n (%) | 21 (61.8%) | 20 (58.8%) | 0.804 |
| Time to defervescence (days) | 3.5 ± 2.2 | 3.1 ± 1.3 | 0.416 |
| Pneumonia[†], n (%) | 2 (5.9%) | 5 (14.7%) | 0.428[a] |
| Hypoxia ($SpO_2$ < 94 %), n (%) | 0 (0.0%) | 2 (5.9%) | 0.493[a] |
| Duration of oxygenation (days) | 0.0 ± 0.0 | 4.0 ± 0.0 | NA |
| Treatments | | | |
| High flow, n (%) | 0 (0.0%) | 0 (0.0%) | NA |

| | | | |
|---|---|---|---|
| Regdanvimab, n (%) | 0 (0.0%) | 2 (5.9%) | 0.493[a] |
| Remdesivir, n (%) | 2 (5.9%) | 3 (8.8%) | >0.999[a] |
| Dexamethasone, n (%) | 4 (11.8%) | 3 (8.8%) | >0.999[a] |
| Antimicrobial agents, n (%) | 4 (11.8%) | 6 (17.7%) | 0.494 |

Patients were classified into two groups according to their booster vaccination status. Data are expressed as mean ± standard deviation or number with percentages. BMI, body mass index; CCI, Charlson comorbidity index; CRP, C-reactive protein; IL, interleukin; BT, body temperature; $SpO_2$, saturation of percutaneous oxygen; NA, not applicable

[a]*p*-value was calculated by Fisher's Exact Test.

[†]Presence of pneumonia was determined using chest radiography.

**Supplementary Table 10. Geometric mean titres of antibodies in patients with breakthrough infection caused by delta and omicron variant**

| | Antibody titre (U/mL) | *p*-value |
|---|---|---|
| Fever | | <0.001 |
| BT ≥ 37.5 ℃ | 1201.5 (715.1-2018.5) | |
| BT < 37.5 ℃ | 4610.4 (3155.5-6736.2) | |
| Hypoxia | | 0.017 |
| $SpO_2$ ≥ 94 % | 98.8 (7.9-1232.2) | |
| $SpO_2$ < 94 % | 2674.5 (1988.0-3598.0) | |
| Pneumonia† | | 0.042 |
| With pneumonia | 774.1 (235.6-2543.3) | |
| Without pneumonia | 2761.0 (2063.6-3694.2) | |
| CRP elevation | | 0.015 |
| CRP > 8 mg/L | 1335.1 (735.8-2422.3) | |
| CRP ≤ 8 mg/L | 3216.0 (2182.4-4739.1) | |
| Lymphopenia | | 0.035 |
| Lymphocyte < 1,100 cell/uL | 1032.2 (420.4-2534.7) | |
| Lymphocyte ≤ 1,100 cell/uL | 2855.5 (2106.2-3871.5) | |

The antibody level was expressed as the geometric mean (95% confidential interval). Abbreviations: BT, body temperature; $SpO_2$, percutaneous oxygen saturation; CRP, C-reactive protein; CCI, Charlson comorbidity index.

†Presence of pneumonia was determined using chest radiography.

### Figure legends

**Supplementary figure 2**. Association of antibody levels and specific symptoms and signs during hospitalisation in fully vaccinated patients with delta or omicron variant breakthrough infection (sensitivity analyses using 3-day threshold)

Antibody levels for each variable representing the clinical course are described as box plots of median and 95% CI in 80 patients with available antibody levels.

**Supplementary figure 3**. Association of antibody levels and specific symptoms and signs during hospitalisation in fully vaccinated patients with delta or omicron variant breakthrough infection (sensitivity analyses using 5-day threshold)

Antibody levels for each variable representing the clinical course are described as box plots of median and 95% CI in 100 patients with available antibody levels.

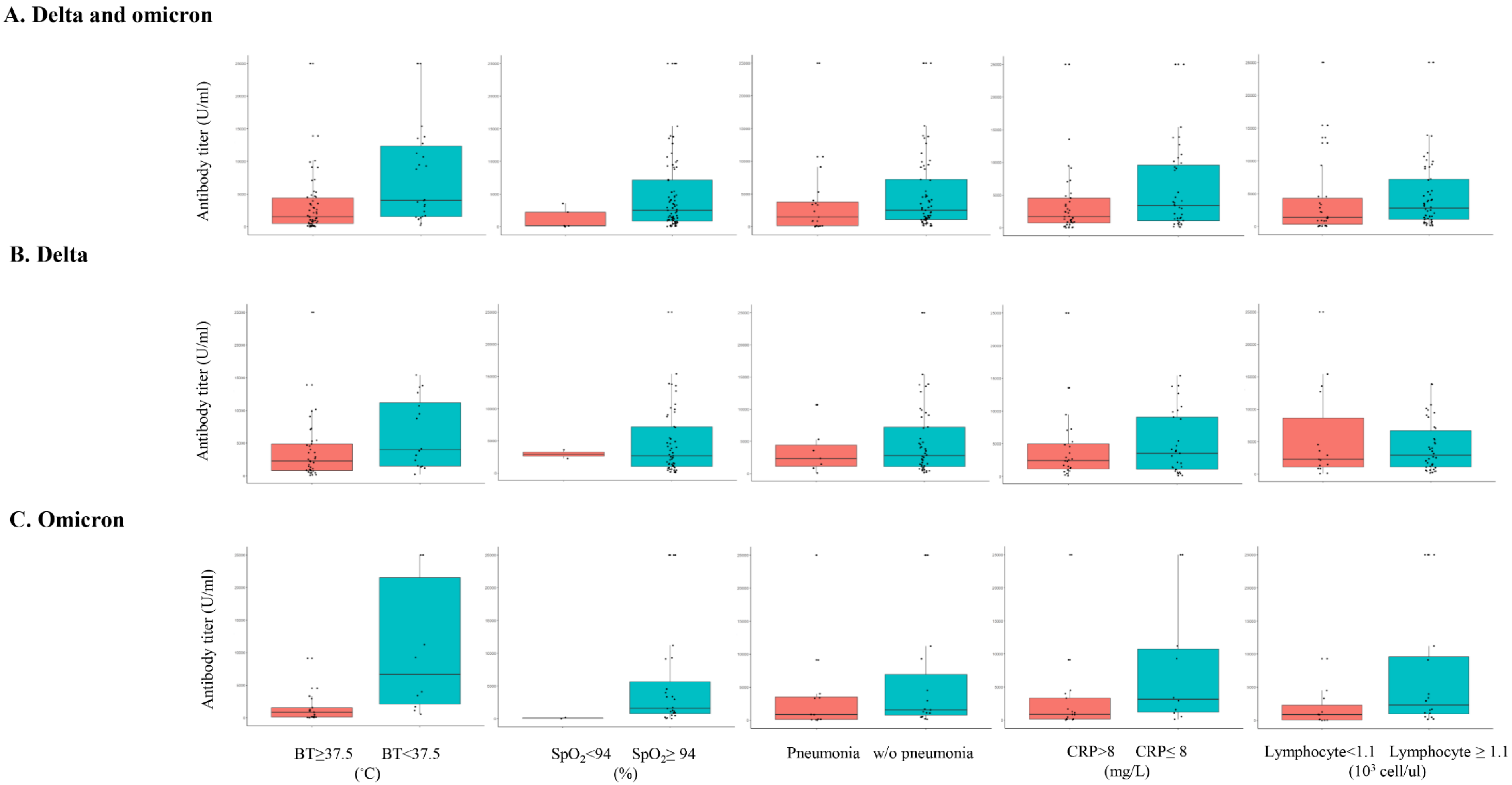


**Supplementary Figure 2.**

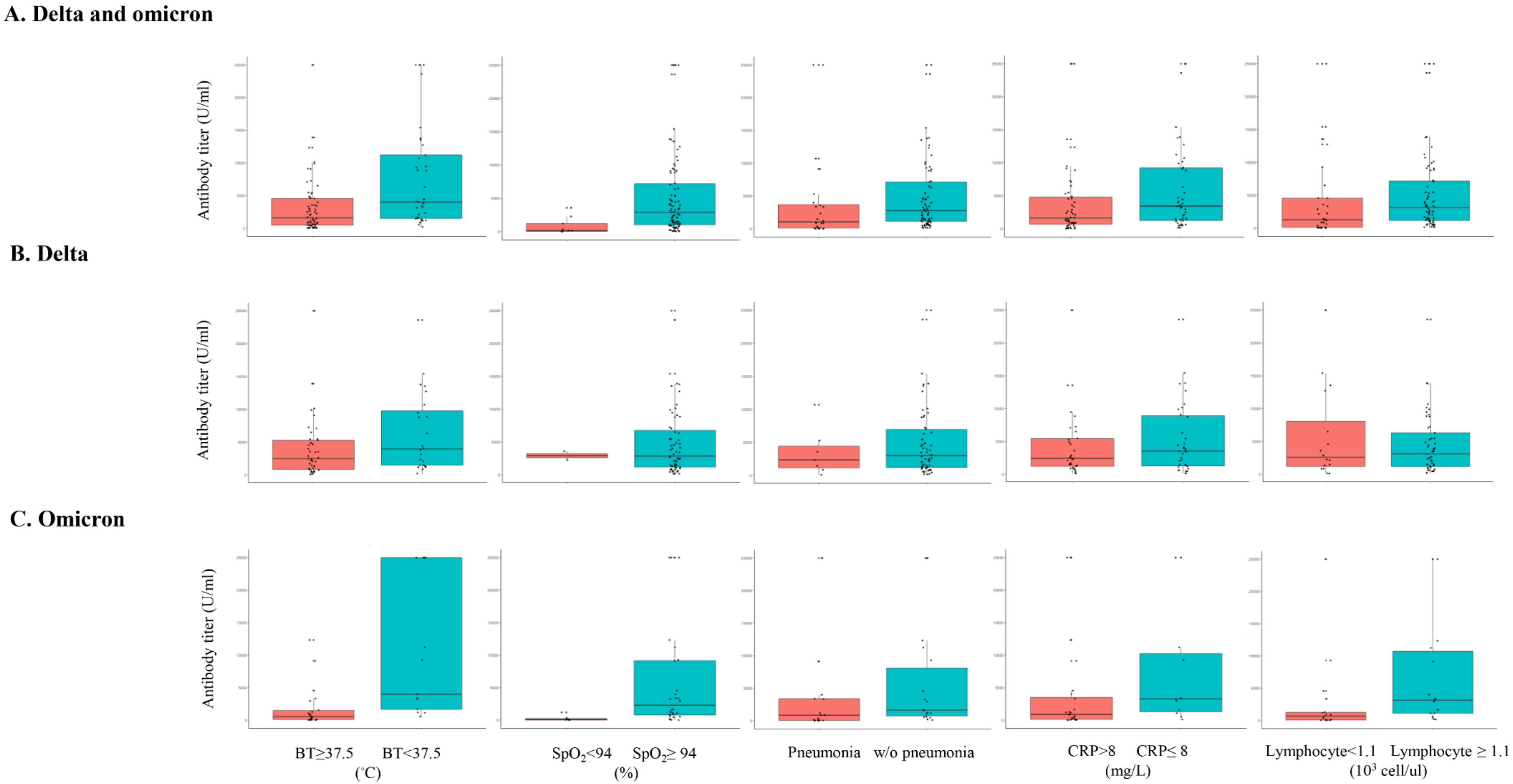


**Supplementary Figure 3.**
